# Ghrelin is associated with an elevated mood after an overnight fast in depression

**DOI:** 10.1101/2023.12.18.23300133

**Authors:** Rauda Fahed, Corinna Schulz, Johannes Klaus, Sabine Ellinger, Martin Walter, Nils B. Kroemer

## Abstract

**Background:** Major depressive disorder (MDD) comprises subtypes with distinct symptom profiles. For example, patients with melancholic and atypical MDD differ in the direction of appetite and body weight changes as well as mood reactivity. Despite reported links to altered energy metabolism, the role of circulating neuropeptides from the gut in modulating such symptoms remains largely elusive.

**Methods:** We collected data from 103 participants, including 51 patients with MDD and 52 healthy control participants (HCP). After an overnight fast, we measured blood levels of (acyl and des-acyl) ghrelin and participants reported their current metabolic and mood states using visual analog scales (VAS). Furthermore, they completed symptom-related questionnaires (i.e., STAI-T).

**Results:** Patients with atypical versus melancholic MDD reported less negative affect (*p* = .025). Higher levels of acyl ghrelin (corrected for BMI) were associated with improved mood (*p* = .012), specifically in patients with MDD. These associations of ghrelin were not mood-item specific and exceeded correlations with trait markers of negative affectivity. In contrast to associations with mood state, higher levels of ghrelin were not associated with increased hunger per se or changes in appetite in patients with MDD.

**Limitations:** The study is limited by the cross-sectional design without an intervention.

**Conclusions:** Our results reveal potentially mood-enhancing effects of ghrelin in fasting individuals that exceed associations with metabolic state ratings. These associations with circulating neuropeptides might help explain anti-depressive effects of fasting interventions and could complement conventional treatments in patients with melancholic MDD.

**Graphical abstract:** 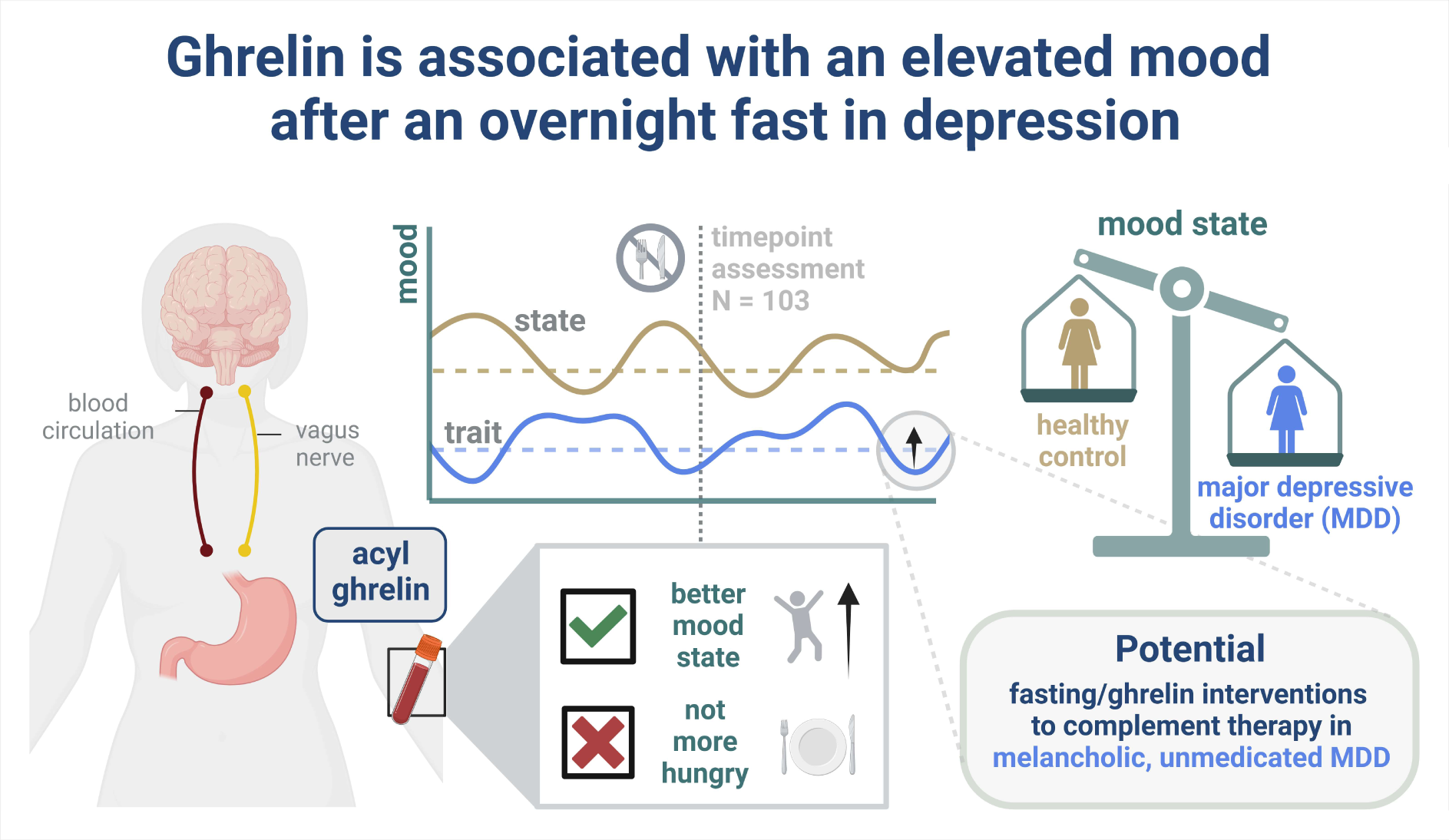

## Introduction

In recent years, fasting interventions, such as intermittent fasting, have gained traction due to reported positive effects on physical health and mental well-being (Patterson and Sears, 2017, Anic et al., 2022). Accordingly, studies testing the effect of fasting on depressive mood in patients with major depressive disorder (MDD) have shown promising results (Berthelot et al., 2021, Fernández-Rodríguez et al., 2022). MDD is a complex and heterogeneous mental disorder characterized by pervasive sadness and negative mood (Uher et al., 2014). In addition to mood disturbances, changes in appetite count toward the diagnostic criteria of MDD (Bosaipo et al., 2017). Whereas the melancholic subtype of MDD experiences a loss of appetite and a pronounced negative affect, the atypical subtype of MDD shows reactive mood fluctuations and an increased appetite (Stewart et al., 2007, Bosaipo et al., 2017). These subtypes of MDD illustrate an intriguing link between disturbances in mood and appetite in depression, but it is not well understood how short-term fasting and corresponding bodily signals that signify metabolic state help regulate mood.

Among the circulating neuropeptides conferring information on metabolic state to the brain, ghrelin has a unique role by facilitating appetitive (Malik et al., 2008). Ghrelin is synthesized in the stomach and when the stomach is empty, more ghrelin is released (Sato et al., 2011). It exists in two distinct forms: acyl ghrelin and des-acyl ghrelin (Hosoda et al., 2000). Initially, acyl ghrelin was regarded as the biologically active form, while des-acyl ghrelin was considered biologically inactive (Delhanty et al., 2012). However, recent research revealed that both acyl and des-acyl ghrelin have significant biological functions (Delhanty et al., 2012, Yanagi et al., 2018, Delhanty et al., 2014). Circulating ghrelin acts primarily on the hypothalamus, which is responsible for maintaining energy balance and appetite regulation (Yanagi et al., 2018). Plasma ghrelin levels rise before eating and decrease post-meal (Cummings et al., 2001), indicating that ghrelin plays a role in meal initiation. In accordance, a recent meta-analysis showed a positive correlation between blood levels of ghrelin and subjective levels of hunger in healthy adults (Anderson et al., 2023).

Although ghrelin is often seen through the lens of energy homeostasis (Gil-Campos et al., 2006), the accumulating evidence on the regulation of goal-directed behavior supports involvement in allostasis. Allostasis is a prospective process through which the brain anticipates and orchestrates physiological adjustments in response to future environmental demands (Katsumi et al., 2022). Its purpose is to facilitate an organism’s long-term survival, growth, movement, and reproduction (Barrett et al., 2016). Metabolic signals, such as ghrelin, therefore convey the brain’s directives to the body (Yanagi et al., 2018), promoting affective states and corresponding instrumental behaviors that align future energy intake with the body’s metabolic needs (MacCormack and Muscatell, 2019, Barrett et al., 2016). Hence, ghrelin’s role may extend beyond sensing of metabolic state by promoting motivational and appetitive behavior (MacCormack and Muscatell, 2019) even before conscious sensations of hunger arise (Frecka and Mattes, 2008). In line with this interpretation, the growth hormone secretagogue receptor type 1a receptor as the primary target of ghrelin is not only expressed in the hypothalamus. Instead, it is widely distributed across the brain, including the hippocampus, substantia nigra, ventral tegmental area, and raphe nuclei (Hattori et al., 2001), and involved in the regulation of a broad range of behavior and cognition, such as memory formation, reward processing, impulsivity, anxiety, and stress vulnerability (Schulz et al., 2023b). Overall, ghrelin plays an important role in promoting appetitive behavior according to metabolic needs through facilitating anticipatory behavior.

In various rodent studies, ghrelin has already been shown to exert anxiolytic, stress-reducing, and antidepressant-like effects by modulating the hypothalamic-pituitary-adrenal (HPA) axis (Lutter et al., 2008, Jensen et al., 2016, Spencer et al., 2012). These findings suggest a protective function of ghrelin against depressive symptoms induced by chronic stress. However, human studies regarding ghrelińs impact on depressive symptoms have reported mixed results to date. Contrary to the association between levels of ghrelin and increased subjective hunger (Anderson et al., 2023), previous studies with patients suffering from MDD revealed that loss of appetite is associated with higher blood levels of ghrelin (Matsuo et al., 2012, Simmons et al., 2020), even after controlling for differences in BMI. Concerning depressed mood, some studies in patients with MDD have reported correlations between ghrelin levels and depressive symptoms (Algul and Ozcelik, 2018, Akter et al., 2014), while other studies found that ghrelin infusions are associated with an improved depressed mood (Kluge et al., 2011). Notwithstanding, other studies observed no association (Kluge et al., 2009, van Andel et al., 2022, Wittekind et al., 2022), adding to the mixed set of interim results. Crucially, most studies did not differentiate between the two isoforms of ghrelin, acyl, and des-acyl ghrelin, which may contribute to the variability in their findings.

To better understand the association of ghrelin with mood in MDD after an overnight fast, we investigated the connection with subjective hunger ratings and contrasted the associations to healthy control participants (HCP). First, we hypothesized that individuals with higher ghrelin levels will report more subjective hunger. Second, we considered atypical symptoms of MDD because this may help resolve inconsistencies in the human literature. We reasoned that patients with MDD and loss of appetite will have lower fasting levels of ghrelin and report being less hungry after fasting compared to healthy control individuals and patients with MDD and an increase in appetite. Third, we predicted that participants with higher fasting levels of ghrelin will report lower levels of negative affect and, potentially, higher levels of positive affect. By advancing the knowledge of ghrelin’s role in mood regulation and its potential involvement in mood disorders, this study may contribute to new targeted therapeutic interventions for MDD.

## Methods

### Participants

For the current analysis, we included data from an ongoing study on the gut-brain axis in depression (https://clinicaltrials.gov/study/NCT05318924; Schulz et al., 2023a). The sample included 103 participants (*M_Age_* = 29.40 ± 7.37, *M_BMI_* = 23.74 ± 3.21 kg/m^2^; Table 1), including 51 patients with MDD, and 52 HCP. To ensure the eligibility of the participants, we conducted phone screenings. Participants were required to be between 20 and 50 years old, with a BMI ranging from 18.5 kg/m² to 30 kg/m². Participants were excluded if they had a history of schizophrenia, bipolar disorder, severe substance abuse, or severe neurological conditions. They were also excluded if they met the criteria for eating disorders, obsessive-compulsive disorder, trauma and stressor-related disorder, or somatic symptom disorder within the last 12 months. Additionally, participants who were on medication or had medical conditions that influenced body weight (except for antidepressants in MDDs), as well as pregnant or nursing women, were excluded. HCPs were selected if they never met the criteria for mood or anxiety disorders, while individuals with MDD had to fulfill the DSM-5 criteria for MDD at the time of participation. In the case of pharmacological treatment, a stable dose of antidepressants for a minimum of two months was required. Before their participation, each participant provided written informed consent. The study was approved by the ethics committee of the Faculty of Medicine at the University of Tübingen. It was conducted in accordance with the Declaration of Helsinki.

**Table 1.** Sample characteristics.

| Characteristic | HCP<br>(N=52) | MDD<br>(N=51) | Overall<br>(N=103) |
| --- | --- | --- | --- |
| **Age [years]** |  |  |  |
| Mean (SD) | 30.46 (± 7.31) | 28.42 (± 7.37) | 29.40 (± 7.37) |
| **BMI [kg/m<sup>2</sup>]** |  |  |  |
| Mean (SD) | 23.81 (± 3.01) | 23.69 (± 3.42) | 23.74 (± 3.21) |
| **Height [m]** |  |  |  |
| Mean (SD) | 1.74 (± 0.088) | 1.72 (± 0.105) | 1.73 (± 0.097) |
| **Waist circumference [cm]** |  |  |  |
| Mean (SD) | 80.79 (± 11.67) | 82.59 (± 11.58) | 81.72 (± 11.59) |
| **Hip circumference [cm]** |  |  |  |
| Mean (SD) | 96.43 (± 13.03) | 96.83 (± 9.84) | 96.64 (± 11.43) |
| **Weight [kg]** |  |  |  |
| Mean (SD) | 72.61 (± 12.11) | 70.84 (± 14.51) | 71.69 (± 13.37) |
| **Acyl ghrelin [pg/mol]** |  |  |  |
| Mean (SD) | 180.60 (± 205.29) | 166.16 (± 212.16) | 173.08 (± 207.92) |
| Missing | 5 (9.6%) | 2 (3.9%) | 7 (6.8%) |
| **Des-acyl ghrelin [pg/mol]** |  |  |  |
| Mean (SD) | 187.66 (± 94.60) | 189.74 (± 113.58) | 188.75 (± 104.38) |
| Missing | 5 (9.6%) | 2 (3.9%) | 7 (6.8%) |
| **BDI** |  |  |  |
| Mean (SD) | 3.39 (± 4.09) | 27.51 (± 9.96) | 16.07 (± 14.35) |
| **STAI-T** |  |  |  |
| Mean (SD) | 34.54 (± 8.44) | 60.12 (± 9.15) | 47.33 (± 15.55) |
| **Anti-depressive medication** |  |  |  |
| None | 52 (100%) | 27 (52.9%) | 79 (76.7%) |
| SSRI | 0 | 13 (25.5%) | 13 (12.6%) |
| Other | 0 | 11 (21.6%) | 11 (10.7%) |

### Experimental procedure

Participants were scheduled for two sessions. Before they visited the laboratory, all participants were asked to complete a comprehensive set of questionnaires to assess various aspects including demographics, eating behavior, physical activity, and symptoms related to depression. To evaluate the symptom severity of MDD we used the Beck Depression Inventory (BDI-II; (Beck et al., 1996)). Trait-like negative affectivity was assessed by the State-Trait Anxiety Inventory (STAI-T; (Spielberger et al., 1983)). Upon arrival for the first session, participants were instructed and signed written informed consent. The first session then included the Structured Clinical Interview for DSM 5 (SCID-5-CV; (First, 2015)), lasting about 0.5 h for HCP and up to 2 h for patients with MDD. Additionally, patients with MDD completed the Structured Interview Guide for the Hamilton Rating Depression Scale with Atypical Depression Supplement (SIGH-ADS; (Williams and Terman, 2003)). HCPs had the option to complete both sessions in a single day, while patients with MDD attended on two separate days.

For the second session, participants arrived at the laboratory after a 12 h overnight fasting period (only unsweetened beverages were allowed). Upon arrival, blood samples were collected to measure levels of ghrelin, glucose, insulin, and triglycerides (Schulz et al., 2023a). Subsequently, we recorded information regarding the participantś last meal and beverage intake, as well as anthropometric measurements including weight, height, waist and hip circumference (Table 1). In women, we also collected information on the menstrual cycle. Participants then commenced a battery of reward-related tasks (Schulz et al., 2023a), which lasted approximately 3.5 h. Throughout the session, they were provided with water ad libitum. To assess their current metabolic and affective state, participants answered state-related questions using a visual analog scale (VAS) after the blood draw (VAS T1) and before the taste test (VAS T2). The session concluded with participants receiving their performance-based compensation (money and food rewards). Upon completion, they were further compensated with either €50 or partial course credit.

### Visual analog scales (VAS)

To assess the participants’ metabolic and affective state, we administered 24 state-related questions, requiring approximately 5 min to complete. For this study, we focus on the ratings of VAS T1, as these were collected directly after we collected blood for the measurement of ghrelin levels. Each question asked participants to rate the items based on their current feelings at that moment. First, participants were asked to assess their current physical and metabolic state. This involved rating their level of hunger, thirst, fullness, and fatigue. Second, to assess their current mood, participants were presented with 20 items of the German version of the Positive and Negative Affect Schedule (Krohne et al., 1996). These items are often divided into two categories: positive affect and negative affect domain (Ferstl et al., 2022, SI1). All items were displayed on a laptop screen, and participants used an Xbox controller joystick to indicate their responses on a visual analog scale (VAS), which ranged from 0, representing “not at all”, to 100, representing “very much”.

### Blood parameters

At the beginning of the behavioral session, we drew blood to measure fasting levels of hormones. Ghrelin samples were obtained using a 9.0 ml K3E-EDTA (anticoagulant) monovette and taken directly to the laboratory to centrifuge the sample at 4°C with 2000 g for 10 min. Then, 500 µl of plasma was pipetted from the EDTA-Monovette into two cooled 1.8 ml cryo bottles (Thermo Scientific™ Nunc™) each and 50 µl of cooled 1 M hydrochloric acid (HCL) in plasma to acid ratio of 10:1 was added to prevent ghrelin from deacetylating. The tubes were immediately capped, gently reversed, and immediately cooled at −20°C before they were relocated to a −80°C freezer (after 24 to 48h). Cryo tubes containing plasma and HCL were analyzed using the acylated ghrelin (human) easy sampling ELISA kit #A05306 and the unacylated ghrelin (human) easy sampling ELISA kit #A05319 from Bertin Bioreagent.

### Data analysis

Ghrelin levels were log-transformed and residualized for age, sex, and BMI. Degrees of freedom were estimated using Satterthwaite approximation and continuous predictors were grand-mean centered. To test for an influence of depression subtype, we calculated the atypical balance score from the SIGH-ADS (Williams and Terman, 2003, SI2).

#### Statistical threshold and software

We considered α < 0.05 as significant. All analyses were conducted with R (v4.3.0; R Core Team 2023).

## Results

### Ghrelin is not associated with subjective metabolic state ratings

To evaluate the association between fasting plasma levels of ghrelin and subjective metabolic state, we calculated the correlation between acyl ghrelin and ratings of hunger, fullness, thirst, and fatigue. Contrary to our hypothesis, neither HCP nor patients with MDD with higher levels of acyl ghrelin reported being hungrier (HCP: *r* = −.06, *p* = .69; MDD: *r* = −.052, *p* = .72; Fig. 1b). Additionally, there were no correlations between fasting levels of acyl ghrelin and ratings of satiety in HCP (*r* = .11, *p* = .48) and patients with MDD (*r* = .14, *p* = .34, Fig. 1c). Similarly, in both groups, no correlations were observed for thirst (HCP: *r* = .075, *p* = .62; MDD: *r* = .035, *p* = .81, Fig. 1d), and fatigue (HCP: *r* = .11, *p* = .48; MDD: *r* = −.22, *p* = .12). Use of antidepressants did not affect the correlations. For des-acyl ghrelin, there were also no significant correlations with metabolic state ratings (Fig. S1). Furthermore, levels of acyl ghrelin were not associated with specific symptoms of atypical MDD, including an increase in appetite (*r* = .09, *p* = .55), increased eating (*r* = .02, *p* = .87), and weight gain (*r* = −.10, *p* = .48). These findings suggest that fasting levels of ghrelin are not associated with the subjective perception of hunger or changes in metabolic state as symptoms of MDD.

**Figure 1.**
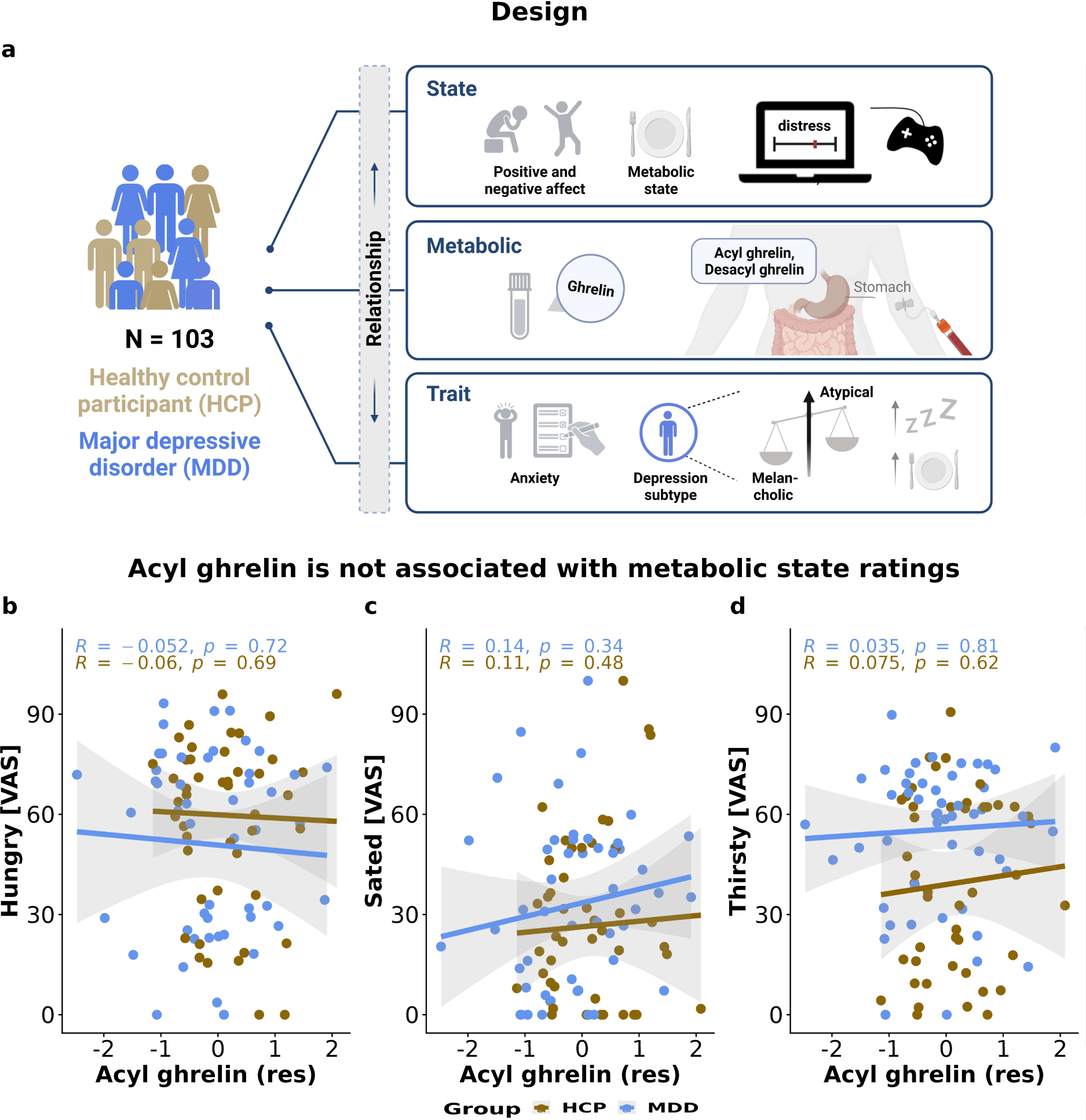
Fasting levels of acyl ghrelin were not associated with metabolic state ratings. A: To investigate the relationship between plasma levels of ghrelin and subjective metabolic and mood state, 103 participants, comprising 52 HCP and 51 patients with MDD, were invited after a 12-hour overnight fast to the laboratory. Following a blood draw, participants indicated their current subjective metabolic state and their mood on 24 Visual analog scales (VAS). Additionally, they completed various questionnaires to assess their trait anxiety and depression subtype. B-D: After log-transforming and residualizing for age, sex, and BMI fasting levels of acyl ghrelin were not associated with subjective ratings of hunger (HCP: *r* = −.06, *p* = .69; MDD: *r* = −.052, *p* = .72), satiety (HCP: *r* = .11, *p* = .48; MDD: *r* = .14, *p* = .34), and thirst (HCP: *r* = .075, *p* = .62; MDD: *r* = .035, *p* = .81) in HCP and MDD. For des-acyl ghrelin there were also no correlations with these metabolic state items (Fig. S1).

### Higher levels of acyl ghrelin are associated with an elevated mood

To investigate the association between ghrelin and mood, we calculated the correlations between blood levels of ghrelin (acyl and des-acyl) and mood state. Mood state reflects the difference between positive mood items and negative mood items. Across groups, higher blood levels of acyl ghrelin were associated with an elevated mood (*r* = .25, *p* = .012, Fig. 2a). This association was driven by patients with MDD (*r* = .29, *p* = .042), while there was no association in HCP alone (*r* = −.052, *p* = .73). We demonstrated that patients with atypical MDD show less negative affect than patients with melancholic MDD (*r* = −.32, *p* = .025; S2). However, as previously mentioned there was no correlation between acyl ghrelin and specific symptoms of atypical MDD. In patients with MDD, the correlation between acyl ghrelin and mood state was slightly larger in patients without any medication (*r* = .33, *p* = .092, Fig. 2d), compared to patients taking SSRIs (*r* = .038, *p* = .90), but not patients using other antidepressants (*r* = .35, *p* = .32). For negative mood alone, acyl ghrelin showed stronger associations in patients without medication (*r* = −.44, *p* = .021, Fig. 2f) in comparison to patients taking SSRI (*r* = .12, *p* = .69) or other antidepressants (*r* = −.17, *p* = .64). However, for positive mood, patients with other antidepressants showed the strongest associations (*r* = .46, *p* = .18, 2e). For des-acyl ghrelin, we observed weaker and non-significant correlations with mood items. This suggests that fasting levels of acyl ghrelin, rather than des-acyl ghrelin, enhance mood, and that this association is attenuated in patients taking SSRIs.

**Figure 2.**
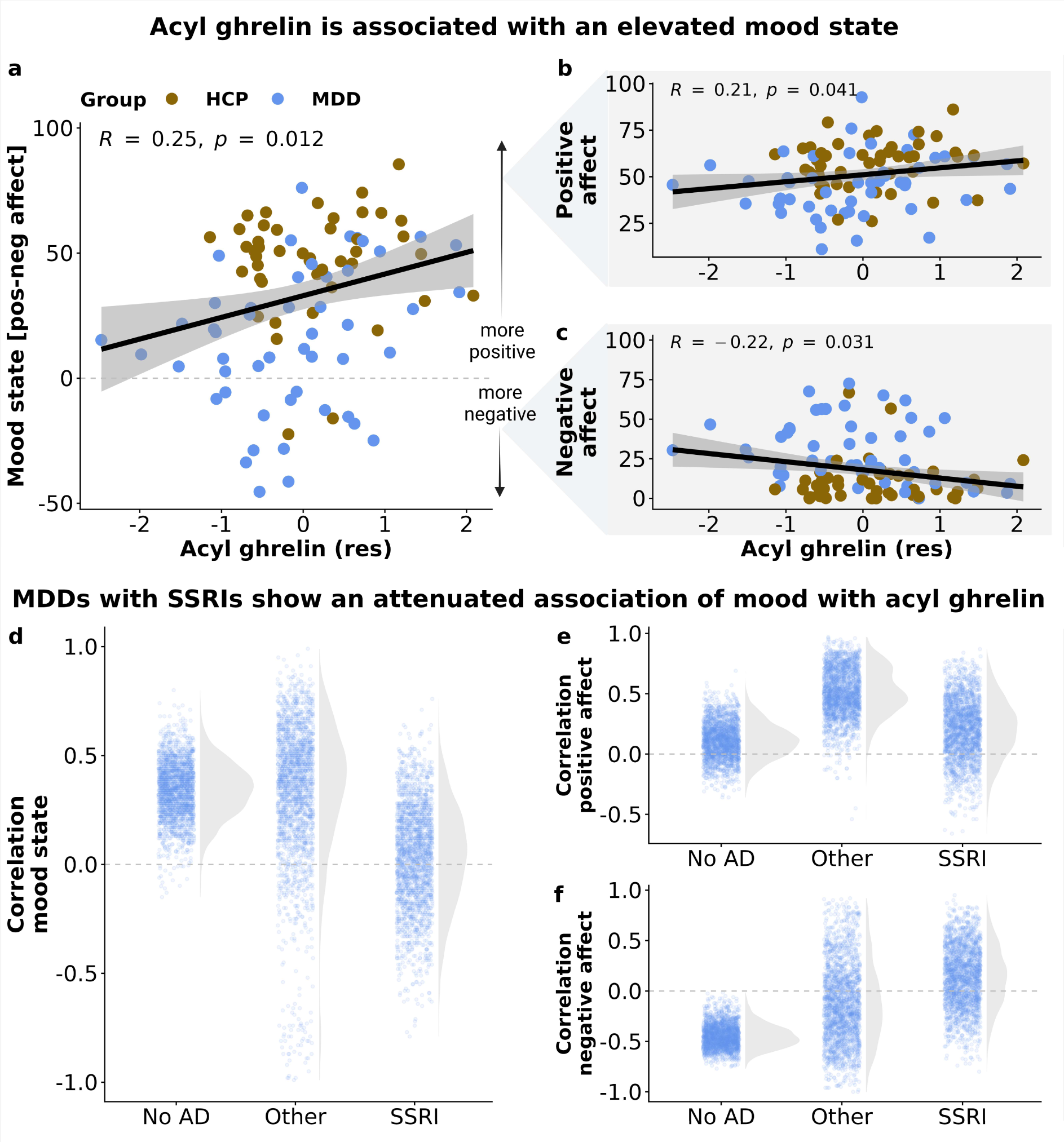
Higher levels of acyl ghrelin are associated with an elevated mood. A-C: Participants with higher levels of acyl ghrelin after fasting show a higher mood state (*r* = .25, *p* = .012), a higher positive affect (*r* = .21, *p* = .041), and less negative affect (*r* = −.22, *p* = .031). D: When focusing on patients with MDD, the correlation between levels of acyl ghrelin and mood state was strongest in patients without any medication (*r* = .33, *p* = .091). Depicted are bootstrapped correlation estimates (*B* = 2000). E: For positive mood alone, we did not find differences across groups. F: For negative mood alone, acyl ghrelin showed stronger associations in patients without medication (*r* = −.44, *p* = .021) in comparison to patients taking SSRI (*r* = .12, *p* = .69) or other antidepressants (*r* = −.17, *p* = .64). No AD = no antidepressive medication, SSRI = selective serotonin reuptake inhibitors, Other = non-SSRI medication.

### The association between acyl ghrelin and mood is not item-specific

Next, we evaluated whether the association of acyl ghrelin is item-specific or more general. In patients with MDD, acyl ghrelin showed the strongest associations with the negative mood item “afraid” (*r* = −.34, *p* = .015, Fig. 3a) and the positive mood item “active” (*r* = .26, *p* = .071, Fig. 3b). To investigate whether acyl ghrelin has more specific effect on facets of mood (e.g., on anxiety), we conducted a factor analysis to extract an unrotated factor that reflects shared variance among all mood items. Then, we compared the correlations between acyl ghrelin and each mood item, both before and after accounting for this factor. This revealed that the link between acyl ghrelin and mood is primarily attributable to the common factor, rather item-specific effect (Fig. 3c). Hence, our analyses point to a general association of acyl ghrelin with enhanced mood.

**Figure 3.**
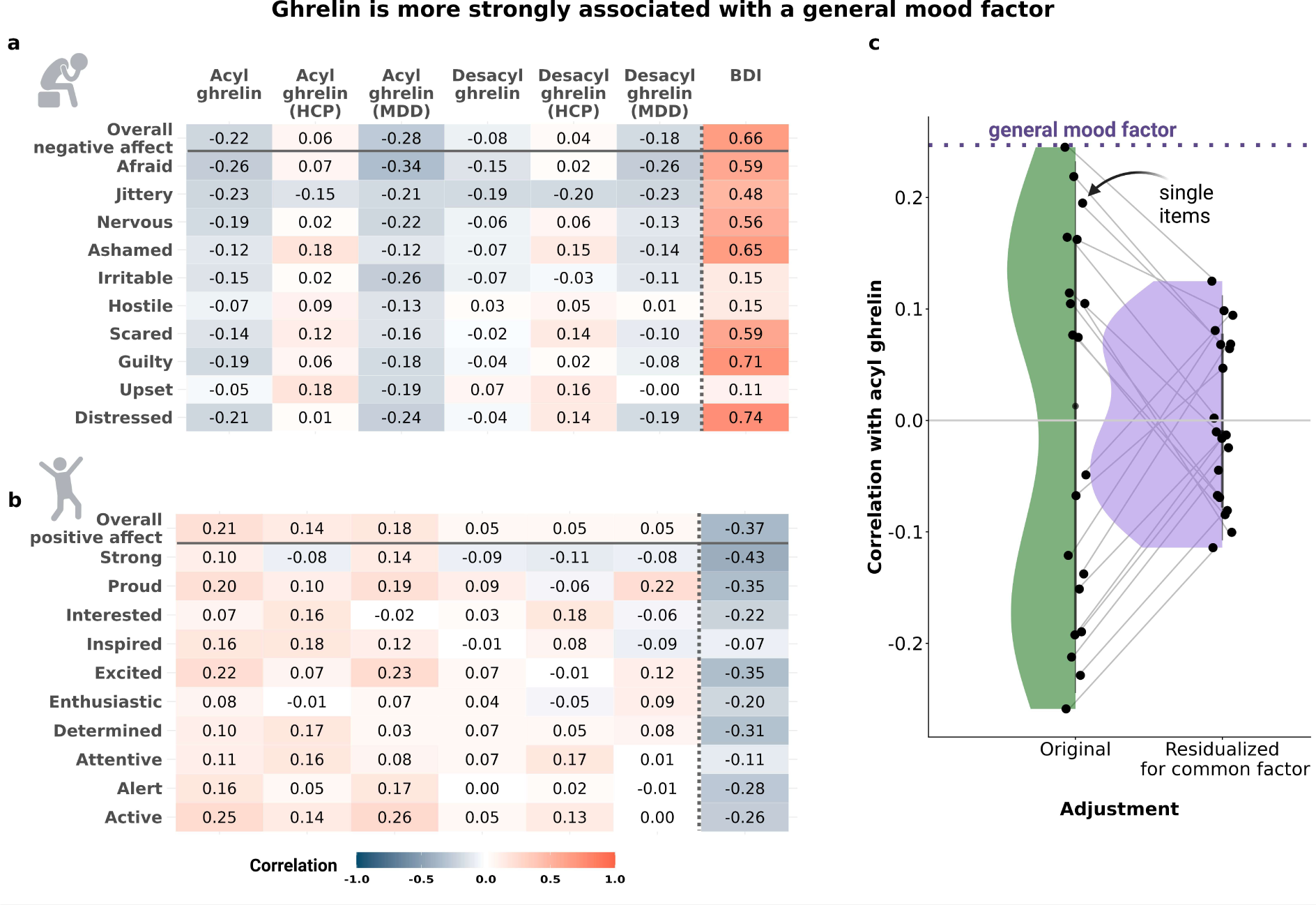
The association between fasting levels of acyl ghrelin and mood is not specific to items, but attributable to overall mood. A: We found negative correlations between levels of acyl ghrelin and ratings across all items of negative affect, particularly in patients with MDD with the strongest association for the item “afraid” (*r* = −.34, *p* = .015). B: There were positive correlations between levels of acyl ghrelin and overall positive affect as well as ratings across all items of positive mood. For patients with MDD, the strongest association was observed for the item “active” (*r* = .26, *p* = .071). C: To investigate if the association between acyl ghrelin and mood is driven by specific items, we extracted a common factor and recalculated the original correlations for each mood item (residualized items). Our analysis shows that the link between acyl ghrelin and mood is not item-specific. The broken line depicts the association with mood state the unrotated factor, demonstrating that single items do not exceed this association.

### Ghrelin is primarily associated with state, not trait-like markers of negative affectivity

Next, after showing an association with mood state ratings, we evaluated if acyl ghrelin is associated with trait-like markers of negative affectivity. As expected, STAI-T scores were correlated with negative mood items (Fig. 4a). Moreover, STAI-T scores were negatively correlated with positive mood items, even though associations were lower overall compared to negative mood items (Fig. 4b). Higher fasting levels of acyl ghrelin were numerically associated with slightly lower STAI-T scores (*r* = −.15, *p* = .15; Fig. 4c), but this association was weaker compared to state mood (*r* = .25, *p* = .012). Furthermore, we did not find correlations between STAI-T scores and hunger ratings after fasting (*r* = −.12, *p* = .22). Hence, acyl ghrelin appears to be more closely associated with the current mood state, even though the momentary expression of negative mood is strongly associated with trait-like dimensions reflecting negative affectivity.

**Figure 4.**
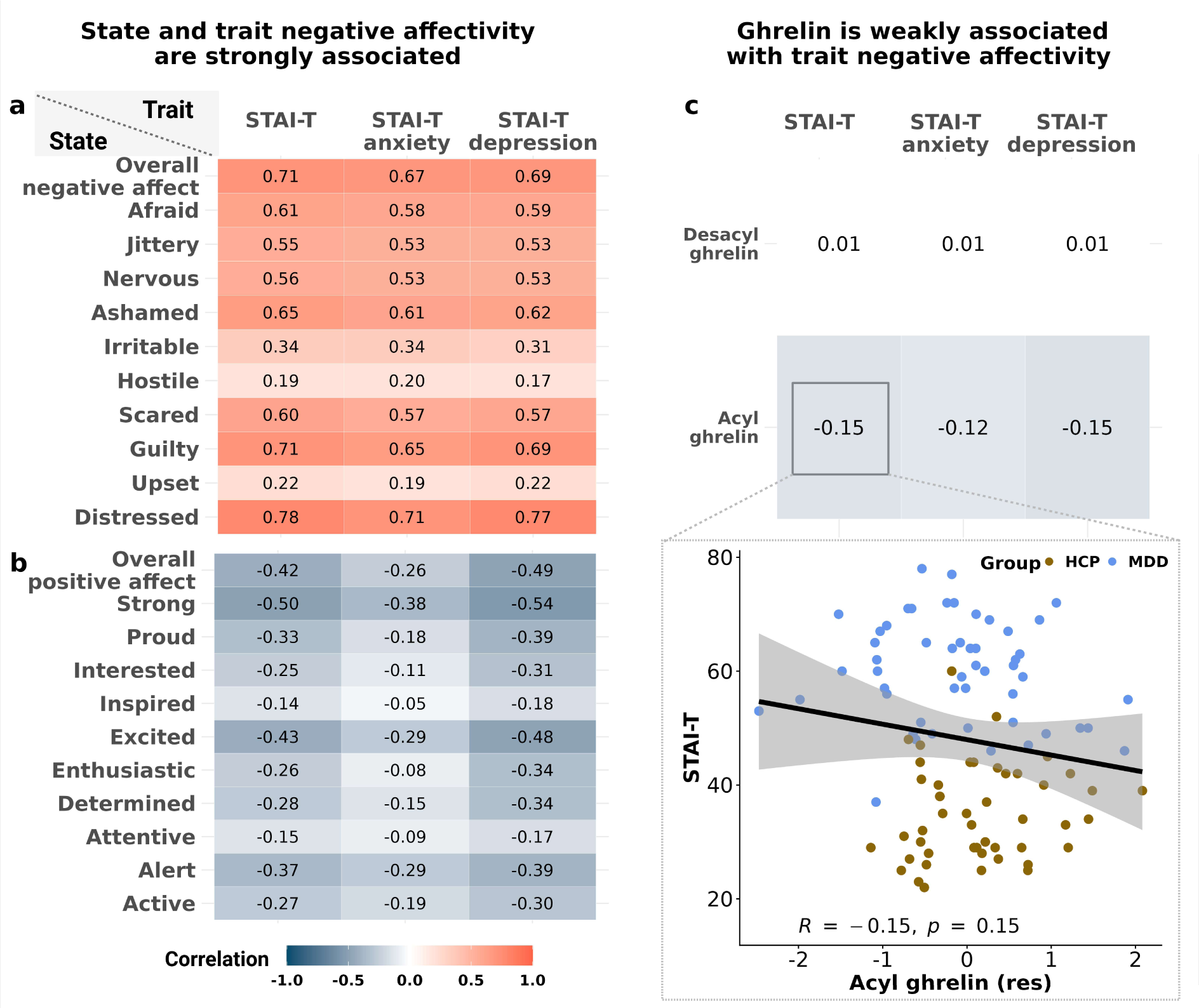
Fasting levels of acyl ghrelin are more strongly associated with state versus trait-like markers of negative affectivity. A: To measure negative affectivity, we calculated the depression and anxiety subscores of the STAI-T. Scores of the STAI-T were strongly correlated with negative mood ratings. B: Scores of the STAI-T were moderately correlated with positive mood ratings. C: Acyl ghrelin was not associated with STAI-T scores (overall STAI-T: *r* = −.15, *p* = .15). No correlations between STAI-T sub-scores and levels of acyl and des-acyl ghrelin.

## Discussion

Depression is characterized by marked changes in appetitive behavior and mood, but modulatory effects of circulating neuropeptides from the gut are still poorly understood. Here, we show that fasting levels of ghrelin exert mood-enhancing effects that exceed associations with subjective ratings of metabolic state, particularly in patients with MDD. Notably, these associations were a) not specific to a few narrow items, b) larger in patients without concurrent antidepressant medication, and c) exceeded correlations with trait-like measures of negative affectivity. Perhaps surprisingly, the associations of ghrelin with mood were largely independent of MDD symptom dimensions, although patients with melancholic MDD reported more negative mood. To conclude, our study shows that ghrelin may play a role in the mood-enhancing effects of fasting, which highlights the potential of signals from the gut to improve key symptoms of depression.

Our finding that fasting levels of ghrelin are associated with an elevated mood adds to the growing literature demonstrating a link between putative homeostatic signals and affective tone. Preclinical studies have shown that ghrelin regulates stress-induced neural and behavioral responses reducing depressive symptoms (Lutter et al., 2008, Kanehisa et al., 2006) and anxiety (Jensen et al., 2016) via the hypothalamic-pituitary-adrenal (HPA) axis (Spencer et al., 2012). However, chronic administration of ghrelin may have detrimental effects on mood and behavioral measures reflecting depressive states in rodents (Hansson et al., 2011). Anxiolytic effects have been shown to be mediated by the basolateral amygdala because local overexpression of the ghrelin receptor increases ghrelin-induced effects (Jensen et al., 2016). In contrast, findings in human studies have been mixed (Spencer et al., 2015) and heterogeneity in symptoms related to appetite and body weight might be one source of inter-individual variability of ghrelin’s action on mood and reward function more broadly (Simmons et al., 2020, Simmons et al., 2016, Kroemer et al., 2022). Our study adds one more dimension that may explain the heterogeneous results in humans since we observed stronger associations in unmedicated patients with MDD. In rats, SSRIs have been shown to exert inhibitory effects on acyl ghrelin and gastrointestinal motor activities through 5-HT2c receptors (Fujitsuka et al., 2009). Comparable effects in humans may counteract the mood-enhancing effect of ghrelin, which might alter the effectiveness of fasting interventions as well. Likewise, metabolic dysregulation before the initiation of an SSRI treatment has been shown to reduce the effectiveness of the medication (Hough et al., 2021), further illustrating the link between energy metabolism, antidepressant medication, and intended effects on mood.

Perhaps surprisingly, we found that fasting levels of ghrelin were more strongly associated with mood compared to metabolic state ratings. However, there are several plausible explanations for a lack of association in our sample. First, we focused on an overnight fasting state when most participants were very hungry. It is possible that this state of homeostatic need masks the role of ghrelin in facilitating appetitive behavior (Malik et al., 2008, Kroemer et al., 2013, Goldstone et al., 2014) if almost everyone is hungry. Second, our findings may highlight the broader function of ghrelin in allostatic behavior. According to a narrow definition of homeostatic function, one may speculate that surges in ghrelin appear in parallel to negative mood states which encourage food-seeking behavior to re-balance the energy budget of the organism (Yanagi et al., 2018, Keramati and Gutkin, 2014). In contrast, the alleged allostatic function is in line with a mood-enhancing effect that promotes food-seeking behavior and approach by tuning reward responsivity (Schulz et al., 2023b). Hence, it is conceivable that mood state shows stronger associations with fasting levels of ghrelin while associations with metabolic state might be larger when participants are neither hungry, nor full.

Despite the clinically relevant demonstration of a mood-enhancing association with higher fasting levels of ghrelin, several limitations will need to be addressed in future studies. First, we only investigated inter-individual differences in endogenous levels of ghrelin. Although it is conceivable that fasting-induced improvements in mood are mechanistically related to marked changes in circulating neuropeptides, ghrelin is only one of the candidate signals of the gut–brain axis (Margolis et al., 2021, Pozzi et al., 2019, Hebb et al., 2005). Consequently, interventional studies with experimental administration of ghrelin are necessary to substantiate the specific link between ghrelin and mood. Second, longer fasting or weight loss interventions could be highly instructive to unravel individual dynamics of mood-related adjustments in correspondence with changes in blood levels of various circulating neuropeptides (Kessler et al., 2018, Neseliler et al., 2019). Third, our study provided preliminary support for larger associations of fasting levels of ghrelin with negative mood in unmedicated patients with MDD, but it was not powered to resolve potential interactions with specific types of antidepressants. Future research may focus on interactions of the gut–brain axis with conventional medications in more detail as there is growing evidence for an interdependence (West et al., 2021, McVey Neufeld et al., 2019) that could ultimately help improve the effectiveness of stratified treatments for MDD.

Circulating neuropeptides from the gut have the potential to improve reward function and mood disturbances, but previous work has yielded inconsistent results in depression. In line with the hypothesized anxiolytic function of ghrelin, we observed mood-enhancing effects of higher fasting levels after a short-term overnight fast, specifically in unmedicated patients with MDD. Notably, these associations of ghrelin with a better mood state generalized across negative and even positive items and exceeded correlations with ratings of metabolic state or trait-like markers of negative affectivity. Taken together, our findings provide support for a broader role of gut-derived signals in regulating mood (Ferstl et al., 2022, Lach et al., 2018, Lutter et al., 2008) and call for more clinical studies to investigate the potential of the gut–axis in rapidly improving key symptoms of depression.

## Acknowledgment

We thank Franziska Peglow, Ebru Sarmisak, Nora Gerth, Yul Wegner, Anne Schiller, Antonia Schlaich, Johanna Voß, and Hannah Groß for help with data acquisition as well as Stephanie Ebbinghaus for support in conducting the ghrelin analyses. Figures were adapted using BioRender.com. The study was supported by DFG KR 4555/7-1, KR 4555/9-1, and KR 4555/10-1, and WA 2673/15-1.

## Author contributions

NBK was responsible for the study concept and design and MW contributed to the conceptualization. RF, CS, & JK collected data under supervision by NBK. RF, CS, & NBK processed the data and performed analyses. SE provided the resources for analyzing blood values. RF, CS, & NBK wrote the manuscript. All authors contributed to the interpretation of findings, provided critical revision of the manuscript for important intellectual content, and approved the final version for publication.

## Data availability statement

All data produced in the present study are available upon request to the authors.

