## Supplemental Material 1 for "Ghrelin is associated with an elevated mood after an overnight fast in depression"

**Supporting information**

**SI1. Overall positive and negative affect**

For positive affect, participants indicated how active, interested, excited, strong, inspired, proud, enthusiastic, alert, determined, and attentive they were feeling. For negative affect, participants reported their levels of distress, upset, guilt, fright, hostility, irritability, shame, nervousness, jitteriness, and fearfulness.

**SI2. Atypical Balance Score**

The total 25-item SIGH-ADS score comprises the 17-item Hamilton score and the 8-item Atypical Symptom score (Williams and Terman, 2003). The Atypcial Symptom score includes the following items: Social withdrawal, weight gain, appetite increase, increased eating, carbohydrate craving or eating, hypersomnia, fatigability, and diurnal variation type B. To calculate the atypical balance score, we divided the 8-item Atypical Symptom score by the total 25-item SIGH-ADS score and multiplied it by 100 (Williams and Terman, 2003). Consequently, the atypical balance score indicates the degree of atypicality, ranging from 0 (minimum) to 100% (maximum).

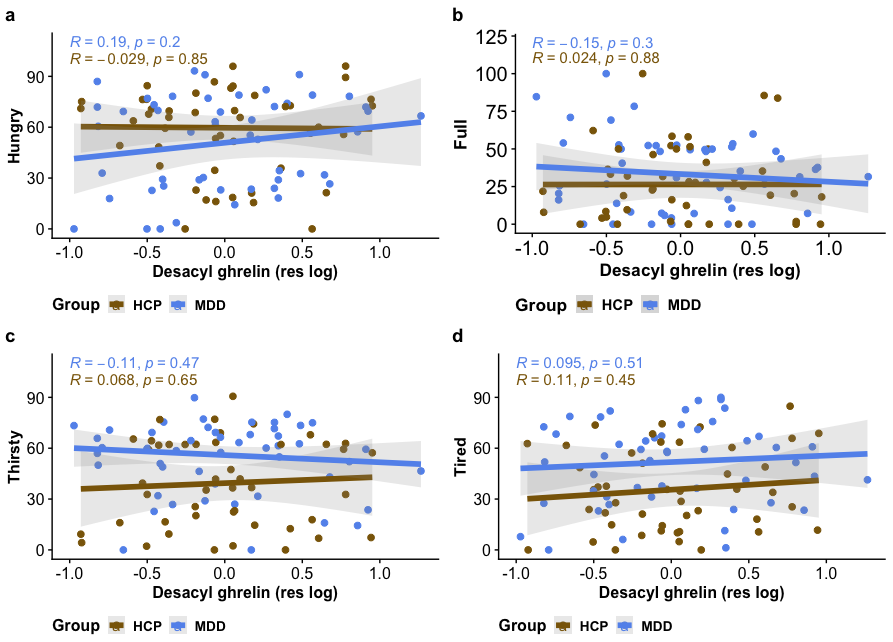

**Fig. S1. No correlation between metabolic state items and levels of des-acyl ghrelin.**

Fasting blood levels of des-acyl ghrelin were not associated with subjective metabolic state ratings in patients with MDD and in HCP. Values for des-acyl ghrelin were log-transformed and residualized for age, sex, and BMI.

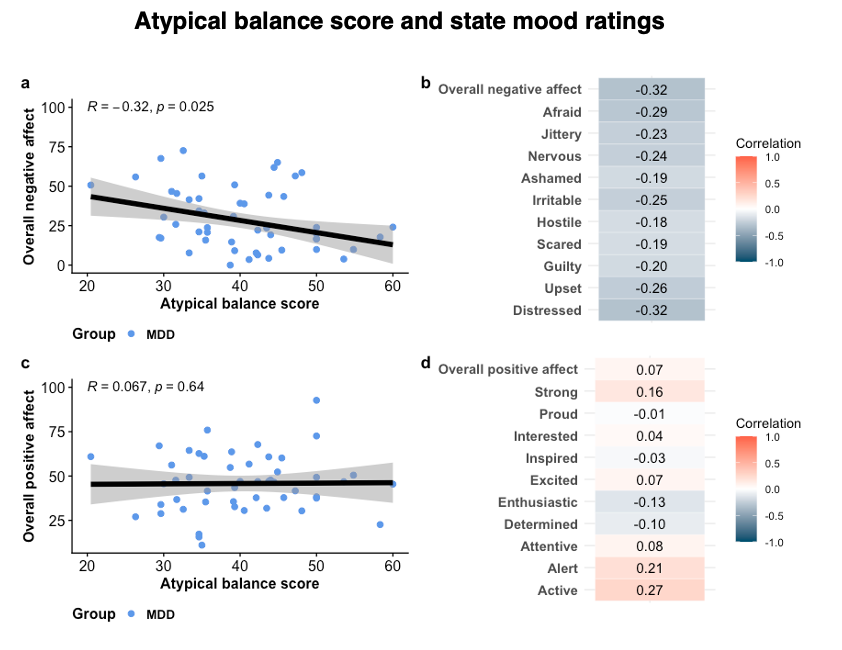

**Fig. S2. Patients with atypical MDD report less negative affect**

We determined whether patients with atypical MDD show differences in the expression of negative mood compared to patients with melancholic MDD. A-B: We found that patients with more atypical symptoms (as indicated by the atypical balance score) showed less negative affect (*r* = -.32, *p* = .025) across all items of negative mood. C-D: For positive mood items, we observed positive correlations of atypical symptoms with the items “active” and “alert”, but not the overall score across items.
